# XTRA Study protocol: eXploring medical sTudents’ caReer reAdiness - A cross sectional study in the UK

**DOI:** 10.1101/2022.01.12.21267945

**Authors:** Amanda Godoi, Mia McDade-Kumar, Farazi Virk, Charlotte Casteleyn, Omar Marei, Ting Yang, Ahmed Moussa, Kashif Aman, Matthew H V Byrne, Priya Rose Babu, Sanya Trikha, Kiana Bamdad, Michal Tombs

**Author notes:** **Corresponding author** Name: Amanda Godoi, Email address, Full Institution address: Cardiff University, School of Medicine, Cardiff, UK, Twitter: @amandagodoimed. **Contact information** Amanda Godoi; Mia McDade-Kumar; Farazi Virk; Charlotte Casteleyn; Omar Marei; Ting Yang; Ahmed Moussa; Kashif Aman; Matthew H V Byrne; Michal Tombs. **Authorship** AG and MT were responsible for conceptualisation. AG, FV, CC, TY, MM, KA, AM were responsible for writing the protocol and first draft. All authors were responsible for writing the second draft. AG, MT and MHVB were responsible for study design. MM, CC and FV were responsible for leading the data collection. All authors were responsible for data collection. The contributions from PB, ST and KB were solely data collection in accordance with collaborative authorship criteria. All authors were responsible for revisions. MT was responsible for supervision. **Study type:** Cross-sectional questionnaire-based study. **Ethical considerations** This study is conducted by Cardiff University, and has been reviewed and approved by the Cardiff University Research Ethics Committee (Reference: SMREC 21/86).

## Abstract

**Background and objectives:** Professional and career enhancing opportunities are essential for developing skills required for a successful career in medicine. Research to date has mainly focused on the extent to which medical schools prepare students for clinical work as junior doctors. However, there remains a need to ascertain how students prepare for their career and what facilitates or hinders learning regarding careers in medicine. The purpose of the XTRA study is to examine career readiness of medical students at UK universities and the support they receive during their studies regarding career planning.

**Methods:** The eXploring medical sTudents’ caReer reAdiness (XTRA) study is a national cross sectional study of all medical students enrolled at a UK medical school. Data collection will occur via a secure online survey designed as a training need analysis based on the principles of Super’s theory (Super, 1953) of career development. A snowball sampling strategy will be used to recruit participants via social media and networks. Results will be analysed using quantitative analysis and thematic analysis to identify themes in qualitative responses. The primary outcome is to understand the perspective of current medical students on how well prepared they are about entering their careers in healthcare.

**Conclusions:** We anticipate that findings from this study will help identify career readiness of medical students to facilitate the development of career development programmes and resources to ensure medical students are well equipped for their future careers.

## Introduction

### Background

Every medical school aims to ensure that all students, at the time of graduating, are ready to start work as medical practitioners. The current literature contains a number of studies that evaluate medical students’ preparedness to practice [1], however, little has been studied about medical students’ career readiness. To have a strong medical career it is important to engage in personal development and seek all available opportunities which are important in improving a practitioner’s portfolio, and developing skills needed to be a doctor [2]. Early consideration of postgraduate career preparation and helpful medical school career guidance has a strong association with perceptions of preparedness of medical graduates for hospital practice [3]. Thus, it is important to gauge how knowledgeable medical students are about career development so that any inadequacies can be addressed. This would help ensure that junior doctors feel prepared and confident about their future, giving them the freedom to make decisions and engage in the best career opportunities for them.

Understanding the requirements for a career in medicine and the opportunities available for a desired trajectory is essential [4]. For example, familiarisation with medical career pathways, networking with other professionals, and understanding the financial aspects of a doctor’s life (such as taxes and pensions) are examples of essential aspects aiding students’ personal development. These factors are crucial to decide the paths students will take [5, 6]. Students will therefore require support on how to develop a professional career in medicine, gaining experiences with teaching and leadership, engaging in research and entrepreneurship, attending conferences and courses, and using social media effectively. Many of these are important components of a medical portfolio, which are needed for applications for training and career progression [7-10].

Having a structured career plan can aid understanding of the skills that need to be developed in a logical time frame, and early career planning has many potential benefits, including better work-life balance [11]. Super’s theory of career development [13, 14] offers some valuable insight into a career coaching structure with three phases: 1) Crystallisation (self-assessment of the medical student’s needs, values, competencies, and opportunities), 2) Specification (establishing preferences regarding specific vocation and career pathways), and 3) Implementation (training and making efforts to establish a specified career goal) [12-14]. This highlights a potential career preparation model that could be relevant to UK medical practitioners, emphasising how career coaching would allow students to develop their career continuously throughout their lives. In our context, the utilization of the Communities of Practice elements as proposed by Lave and Wenger [15, 16] associated with the concept of Hidden Curriculum as proposed by Hafferty and Franks [17, 18] can also offer a powerful tool to explore and understand how medical schools, role models/mentors and medical communities are influencing personal and professional development of medical practitioners..

### Purpose

There is ample evidence to suggest that supporting medical students’ career development can benefit them individually, as well as the medical profession as a whole [2, 4, 8, 10, 11]. Given the importance of such attributes, there is a need to evaluate how students are learning about the process of career readiness and which resources are facilitating or hindering their learning regarding careers in medicine. The XTRA study is therefore a needs assessment and to this end it sets to examine the extent to which students who are currently studying at UK universities feel ready for their career and the opportunities they have to enhance knowledge about career planning [19]. Findings will help formalise, highlight, and establish the basis of subsequent planning, as well as examine whether there is a need to address these requirements in more formal methods to form a routine part of training.

### Research questions

The XTRA study will address three main questions:

1. To what extent do current medical students feel ready about entering their careers in healthcare?
2. Which facilitating factors and barriers can influence medical students’ readiness?
3. Are there sufficient resources provided by medical schools, external training and mentoring to prepare medical students for different stages in their careers?

### Outcome measures

#### i. Primary objective

- To evaluate the perspective of current medical students on how well prepared they are about managing and developing their careers as medical practitioners.

#### ii. Secondary objectives

To identify:

- Factors that have facilitated medical students’ development in their careers.
- Barriers medical students face towards developing their careers.

To investigate:

- The extent to which medical students have had access to support and resources provided by medical schools, external training and mentoring in order to prepare for the different stages in their careers.

To aid:

- Educational planning to improve students’ readiness to enter the medical profession.

### Anticipated Outcomes

We anticipate that this study will aid in recognising areas that medical students would like further support on, which will help in the development of future career development programs and resources.

## Methods

The XTRA study is a cross-sectional questionnaire-based study.

### Study Population

Medical students, who are defined as students currently studying medicine or on a pre-clinical/foundation part of a medical degree where a medical degree is guaranteed upon successful completion of their degree.

#### i. Inclusion Criteria

- All UK or international medical students currently enrolled at a UK Medical School.
- An International student is defined in this study using the definition of the UK Council for International Student Affairs (UKCISA) as Non-British students (full-time or part-time in education); OR students whose normal residency is not in the UK and are regarded as students with Overseas/International fee status

#### ii. Exclusion Criteria

- Participants who are not fluent in the English Language.
- Non-medical students
- International medical students who are not studying
- Medical students from a university that is not recognised by the GMC

### Data collection

A poster will be shared on social media platforms (including Facebook, LinkedIn, Instagram and the company website) to reach medical students from each UK medical school. This poster will explicitly express the need for medical students to partake in the study including a link to the online form for participation. The survey will also be shared through personal contacts. Participants will only be allowed to complete the form once to avoid multiple submissions. Once the questionnaire is completed, participants will also be given the option to invite three medical student colleagues to participate in the study (via a snowball approach).

### Survey

A secure online questionnaire has been created and hosted via the Microsoft Office Forms database that is supported by Medical Education from the School of Medicine at Cardiff University [20].

The questionnaire consists of 20 questions. Participants will be asked for their age and gender, the stage of medical training they are in and their respective universities to capture sample characteristics information. The focus will be on asking participants their opinions on how much they have been supported throughout medical school in several areas of professional development. This will then be followed by their opinions on which resources could aid them into improving their preparation for medical professions.

The first part of the questionnaire has been developed by the researchers based on the principles of Super’s theory of career development [13, 14]: it is divided into 1) Crystallization (questions about the participant’s needs, values and competencies), 2) Specification (questions regarding career pathways, as well as administrative and financial aspects relating to a medical career), and 3) Implementation (questions about the skills and efforts they have been making towards the career of a doctor). A focused review of the literature has been conducted to establish which factors asked in each of the sections mentioned are beneficial to a doctor’s career. For the Implementation section in particular, questions have been designed to compare the participant’s knowledge on specific areas and how well they have implemented this into achievements. This can provide useful insights into whether the perceptions and awareness of career readiness is linked to engagement in actions contributing to the development of a medical career.

The second part of the questionnaire focused on the guidance these students have had from multiple resources (formally and informally) as well as facilitating and hindering factors that impacted their level of readiness. This can allow better understanding of the extent of career support received by medical students and suggest future avenues for improvement.

A pilot study of 18 students was performed to ensure questions were unambiguous, and focus group discussions and a cognitive interview was conducted to construct face validity within the questionnaire.

### Data Analysis

This study will follow the STROBE cross-sectional study guidelines [21]. Data will be analysed through quantitative and qualitative methods. Factor analysis and coefficient alpha will be used as statistical methods to analyse the validity and reliability of the newly generated questionnaire. Descriptive statistics and quantitative data will be analysed through descriptive analysis and through correlations. We will qualitatively analyse data using thematic analysis to identify themes within the free text responses [22].

### Authorship

Authorship guidelines from the National Research Collaborative will be followed on all research outcomes (National Research Collaborative & Association of Surgeons in Training Collaborative Consensus Group, 2018). The main researchers of the steering group will be listed as co-authors. There is a mandatory requirement for all co-authors to have an ORCID ID. Provided minimum requirements for authorship are met, individuals involved in the recruitment of participants will be listed as collaborators in the study. The criteria for collaborative authorship are to disseminate the survey at least once weekly through the various methods mentioned in this protocol, and to also have an ORCID ID.

### Expected Outputs

We anticipate that the results of this study will be published in a peer reviewed medical or scientific journal. We will submit the findings of this project using posters and oral presentations to national and international conferences.

## Data Availability

All data produced in the present study are available upon reasonable request to the authors

### Appendix 1 – Survey

Appendix 1 - Survey
| Question | Answer options |
| --- | --- |
| **Demographics and Consent** |  |
| Consent*<br>Participation is completely **voluntary**. Any responses you provide are **anonymous**. By filling this questionnaire, you are giving your consent to participate in this study and for use of your anonymised data in future publications. | MULTIPLE CHOICE: yes; no (if the answer is no, participants will not be able to proceed with the questionnaire) |
| Please read the attached consent form with all the consent statements. |  |
| Do you give your consent for participation in accordance with the consent form? |  |
| I confirm I am a medical student from a UK medical school* | MULTIPLE CHOICE: yes; no (if the answer is no, participants will not be able to proceed with the questionnaire) |
| Age | MULTIPLE CHOICE: Younger than 18 years old; 18 - 21 years old; 22 - 25 years old; 26 - 30 years old; Older than 30 years old |
| Gender | MULTIPLE CHOICE: male, female, other, prefer not to say |
| Are you a graduate entry student on a four-year medical course? | MULTIPLE CHOICE: yes; no |
| Which year in medical school/stage of medical training are you in? | DROP DOWN: year 1, year 2, year 3, year 4, year 5, intercalation, other (please specify) |
| Which University do you currently attend as a medical student? | DROP DOWN: all UK medical schools & prefer not to say |
| **Crystallisation** |  |
| The following questions are about your **personal development, values and life goals**. Please rate them relating to your self-assessment of the aspects below. |  |
| I have fully reflected on my personal attributes as a professional | LIKERT: strongly disagree, disagree, neutral, agree and strongly agree |
| I have thoroughly defined my life goals | LIKERT: strongly disagree, disagree, neutral, agree and strongly agree |
| I have clearly defined my values and passions | LIKERT: strongly disagree, disagree, neutral, agree and strongly agree |
| I have fully understood my strengths and weaknesses | LIKERT: strongly disagree, disagree, neutral, agree and strongly agree |
| I have thoroughly envisioned how my career might look like in 5+ years time | LIKERT: strongly disagree, disagree, neutral, agree and strongly agree |
| **Specification** |  |
| The following questions are about your knowledge on **career pathways and aspects of a** |  |
| **doctor's life.** Please rate them relating to your self assessment of the aspects below. |  |
| I have fully familiarised myself with foundation training pathways | LIKERT: strongly disagree, disagree, neutral, agree and strongly agree |
| I have fully familiarised myself with specialty training pathways | LIKERT: strongly disagree, disagree, neutral, agree and strongly agree |
| I clearly understand the financial aspects of a doctor's career (taxes, pensions and investments) | LIKERT: strongly disagree, disagree, neutral, agree and strongly agree |
| I clearly understand a doctor's working rota and employee entitlements. | LIKERT: strongly disagree, disagree, neutral, agree and strongly agree |
| I clearly understand how to network with other medical professionals | LIKERT: strongly disagree, disagree, neutral, agree and strongly agree |
| I fully understand the skills I need to develop as a doctor | LIKERT: strongly disagree, disagree, neutral, agree and strongly agree |
| **Implementation** |  |
| How well do you understand the components of a strong medical portfolio? | RATING: 1=very poor understanding; 3 = satisfactory understanding; 5 =excellent understanding |
| Please rate the options below in accordance to the level of **knowledge** you have on each of the experiences mentioned.<br>No knowledge = you have no idea how to obtain this experience/skill<br>Little knowledge = you are aware but still uncertain in the process of obtaining this experience/skill<br>Neutral = you are not sure<br>Some knowledge = you know about the process of obtaining this experience/skill but still has a few lacks in understanding<br>A lot of knowledge = you fully understand about the process of obtaining this experience/skill |  |
| How to engage in research | LIKERT: no knowledge, little knowledge, neutral, some knowledge, a lot of knowledge. |
| How to gain teaching experience | LIKERT: no knowledge, little knowledge, neutral, some knowledge, a lot of |
|  | knowledge. |
| How to obtain leadership experience | LIKERT: no knowledge, little knowledge, neutral, some knowledge, a lot of knowledge. |
| How to obtain an additional degree (BSc, MSc, PGCE, PhD...) | LIKERT: no knowledge, little knowledge, neutral, some knowledge, a lot of knowledge. |
| How to pursue opportunities in entrepreneurship | LIKERT: no knowledge, little knowledge, neutral, some knowledge, a lot of knowledge. |
| How to attend courses and conferences | LIKERT: no knowledge, little knowledge, neutral, some knowledge, a lot of knowledge. |
| How to obtain a prize or award | LIKERT: no knowledge, little knowledge, neutral, some knowledge, a lot of knowledge. |
| How to get involved in a quality improvement project | LIKERT: no knowledge, little knowledge, neutral, some knowledge, a lot of knowledge. |
| How to use social media effectively as a doctor | LIKERT: no knowledge, little knowledge, neutral, some knowledge, a lot of knowledge. |
| Please rate the options below in accordance with the experiences **you have gained or accomplished** to date.<br>No experience = you have not had neither have planned this experience or skill<br>Little experience = you have a concrete plan to acquire this experience of skill<br>Some experience = you are in the process of acquiring the experience/skill<br>Moderate experience = you have acquired the experience/skill in 1 occasion<br>A lot of experience = you have acquired the experience/skill in 2 or more occasions |  |
| Authorship in a PubMed-cited study | LIKERT: no experience, little experience, some experience, moderate experience, a lot of experience. |
| Teaching experience | LIKERT: no experience, little experience, some experience, moderate experience, a lot of experience. |
| Leadership experience | LIKERT: no experience, little experience, some experience, moderate experience, a lot of experience. |
| Additional degree (BSc, MSc, PGCE, PhD...) | LIKERT: no experience, little experience, some experience, moderate experience, a lot of experience. |
| Entrepreneurship | LIKERT: no experience, little experience, some experience, moderate experience, a lot of experience. |
| Courses or conferences | LIKERT: no experience, little experience, some experience, moderate experience, a lot of experience. |
| Prizes / distinctions / merits | LIKERT: no experience, little experience, some experience, moderate experience, a lot of experience. |
| Quality improvement projects | LIKERT: no experience, little experience, some experience, moderate experience, a lot of experience. |
| Effective use of social media | LIKERT: no experience, little experience, some experience, moderate experience, a lot of experience. |
| **Extent of support** |  |
| Are you the first person to study medicine at university in your family? | MULTIPLE CHOICE: Yes/no/Prefer not to say |
| To what extent have you received support from the following sources regarding your career development? |  |
| Medical student colleagues (seniors) | LIKERT: never, rarely, occasionally, often, always |
| Medical student colleagues (same year or younger) | LIKERT: never, rarely, occasionally, often, always |
| Senior medical professionals (registrars or consultants) | LIKERT: never, rarely, occasionally, often, always |
| Senior medical professionals (foundation year doctors) | LIKERT: never, rarely, occasionally, often, always |
| Medical school staff | LIKERT: never, rarely, occasionally, often, always |
| Committees or societies | LIKERT: never, rarely, occasionally, often, |
|  | always |
| Mentor | LIKERT: never, rarely, occasionally, often, always |
| Online community of medical professionals | LIKERT: never, rarely, occasionally, often, always |
| Can you identify any **facilitating** factors that have helped you prepare for your future career? If so, can you describe them please.<br>*These can be any resources or anything that you have found useful in your development of personal values and needs, career pathways, and portfolio experiences.* | FREE TEXT |
| Can you identify any possible **barriers** and **challenges** that may have impacted your preparedness for your career? If so, can you describe them please?<br>*These can be any lack of resources or anything that you have found hindering in your development of personal values and needs, career pathways, and portfolio experiences.* | FREE TEXT |
| Do you feel prepared for your **professional career development**?<br>*This includes a clear understanding of your personal values and needs, career pathways, the life of a doctor and having a strong medical portfolio.* | LIKERT: I feel prepared, I feel somewhat prepared, neutral, I feel somewhat unprepared, I feel unprepared |
| **Understanding preparedness in career development** |  |
| Why do you feel prepared or unprepared for your professional career? | FREE TEXT |
| Please let us know about existing or non-existing **resources** that have supported you in your career development? | FREE TEXT |
| Which resource **format(s)** would have enabled you to feel more prepared for your medical career? | CHECKLIST:<br>Weekly/monthly seminars or workshops<br>A one off teaching session with all the |
|  | necessary information<br>One to one support with a mentor<br>Videos guiding you on each topic<br>Direct support from the medical school<br>A website with the necessary information<br>An online community of medical professionals<br>Mobile apps with supporting materials<br>Other |
| Please let us know which format would help you if you chose "Other" in the previous question | FREE TEXT |
| Have you ever provided guidance or support in the careers of your colleagues or younger students? | MULTIPLE CHOICE: Yes/no |
| **End of Survey and Follow Up** |  |
| **We really appreciate your time and effort in completing the questionnaire.**<br>An optional certificate is available via this link:<br>You may also enter an optional prize draw to win a spot in a mentoring programme by entering this form:<br>**<u>We ask you to help us disseminate this survey by recruiting 3 additional medical students to participate in this study</u>** |  |
| Please enter your email address if you would be willing to be contacted for a future study.<br>We may conduct a follow up study to understand in greater depth responses from this study. This would either be by a focus group discussion with a researcher and other survey participants, by a semi-structured interview with a researcher, or an additional survey. By entering your email address, it does not commit you to participating in a future study.<br>Your email address will be separated from |  |
| the data, will not be shared outside the data collection team, and will be deleted in 2 years time. |  |

## Notes

**Sources of support:** Nil

**Conflicts of interests:** None declared

### Competing Interest Statement

The authors have declared no competing interest.

### Funding Statement

This study did not receive any funding

### Author Declarations

This project has been approved by the research committee in the School of Medicine at Cardiff University (Reference: SMREC 21/86).

